# Alphafold, Foldseek and MD in NOTCH3 variants: a cohort study

**DOI:** 10.64898/2026.02.23.26346941

**Authors:** Xuejiao Men, Li Zhang, Sanxin Liu, Shunzhou Wan, Wei Qiu, Zhengqi Lu, Qingfen Yu

**Affiliations:** Department of Neurology, The Third Affiliated Hospital of Sun Yat-sen University; Center for Computational Biophysics and Bioinformatics, The Third Affiliated Hospital of Sun Yat-sen University; Centre for Computational Science, Department of Chemistry, University College London

## Abstract

**Background and Objectives:** *Notch homolog 3 (NOTCH3)* gene variants were fully penetrant to produce the disease phenotype of CADASIL. Aberrant NOTCH3 protein leads to degeneration of vascular SMCs and pericytes, targeting microcirculation dysfunction and blood-brain barrier (BBB) leakage.

**Methods:** We evaluated neuroimaging data of forty patients with *NOTCH3* gene variants including eighteen missense/insertion mutations in epidermal growth factor repeat (EGF), negative regulatory region (NRR), and disordered region (Dis). We performed an AI-driven pipeline integrating AlphaFold3, Foldseek, and molecular dynamics simulations to elucidate clinical and molecular consequences.

**Results:** Distinct domain mutations exhibited characteristic patterns: EGFs 1, 2, 13-15, 32 and Dis correlated with microbleeds/macro-bleeds, lacunes, perivascular spaces, and acute cerebral microinfarcts; EGFs 2, 3, 13-15, 25 with disrupted disulfide bonds or binding motif of protein O-glucosyltransferase 1 (POGLUT1) were predicted to undergo greater structural and functional deteriorations in Notch signaling pathways. NRR/Fab (antigen-binding fragment) destabilized dominant motions and single apo-Dis exhibited low structural disorder. Agreement between computational and experimental data for wild-type EGFs/POGLUT1 and C49F, R75Q, R141C mutants suggests testable hypotheses that advance understanding of cerebral small-vessel disease.

**Discussion:** Targeting POGLUT1 to modulate EGF-like domains and using the Fab region to stabilize NRR complexes may be a promising therapeutic approach deserving rigorous exploration.

## Introduction

*Notch homolog 3 (NOTCH3)* gene variants are a key diagnostic criterion of cerebral autosomal dominant arteriopathy with subcortical infarcts and leukoencephalopathy (CADASIL), which represents the most common heritable form of cerebral small vessel disease (SVD)^1^. *NOTCH3* gene variants have a pathogenic high incidence rate of 3.4/1000 worldwide^2^. However, reports have revealed that CADASIL occurs at a frequency of 2-5/100000 in the general population^3-5^. That is to say, the frequency of *NOTCH3* gene variants is 100-fold higher than expected based on estimates of CADASIL prevalence. Nevertheless, it was regarded that *NOTCH3* gene variants were fully penetrant to produce the disease phenotype^6^. Therefore, *NOTCH3* gene variants may more frequently cause a much milder phenotype, which may even go clinically underdiagnosed.

The *NOTCH3* gene encodes a single-pass transmembrane surface receptor mainly expressed in arterial vascular smooth muscle cells (VSMCs) and capillary pericytes^7^. The pathogenic *NOTCH3* gene variants (over 350 reported) alter the cysteine residue number within epidermal growth factor-like (EGF-like) repeat domains of NOTCH3 extracellular domain (N3ECD)^8, 9^. This fundamental biochemical aberration leads to unpaired cysteines, misfolded proteins, and even aberrant multimerizations. The accumulation of pathological aggregated N3ECD is intrinsically linked to granular osmiophilic material (GOM) deposits within the basal lamina of arterial VSMCs, detectable by electron microscopy in both cerebral and systemic vessels, including skin biopsies^10^. This pathological process is a hallmark and results in degeneration and loss of VSMCs and pericytes in the brain, targeting microcirculation dysfunction and blood-brain barrier (BBB) leakage, which can cause ischaemia and inflammatory infiltration in CADASIL^1, 11, 12^.

On neuroimaging, brain magnetic resonance imaging (MRI) of *NOTCH3* gene variants typically reveals multiple lacunar infarcts and confluent white matter hyperintensities (WMHs), particularly in the anterior temporal lobes, external capsule, and periventricular regions^13^.Cerebral microbleeds and brain atrophy may also be observed in advanced stages of *NOTCH3* gene variants. Clinically, *NOTCH3* gene variants were characterized by recurrent ischemic strokes, migraine with aura, mood disturbances, and progressive cognitive decline, which often leading to vascular dementia in patients^14, 15^.

Nevertheless, there is currently no definitive treatment for CADASIL including gene therapy. The causes of *NOTCH3* gene variants and single protein mutations for the pathogenesis of CADASIL are well known and widely studied, especially for NOTCH3 EGF regions since 1996^16-20^. The key pathological process of CADASIL is NOTCH3 protein aggregation, which can lead to abnormalities in the vascular matrisome^15, 21, 22^. Therefore, NOTCH3 protein-targeting monoclonal antibodies may be a promising therapy for patients with *NOTCH3* gene variants. However, very few studies have focused on the complex protein mutations and the associated protein-protein interactions for the NOTCH3 signaling pathways. The studies for the non-EGFs regions, such as negative regulatory region (NRR) and disordered region (Dis), are extremely insufficient. The precise molecular cascades connecting this genetic defect to characteristic protein alterations (including EGF and non-EGF regions), vascular pathology, and clinical symptoms remain unclear. Given its relatively high incidence rate of 3.4/1000 and the absence of etiological treatment options, research in this area remains of paramount importance^15^.

Between 2018 and 2023, we collected clinical and neuroimaging data from forty patients with genetically confirmed *NOTCH3* gene variants. The study aimed to elucidate the molecular and vascular mechanisms driving disease expression by correlating clinical and imaging features with the predicted structural and functional consequences of specific *NOTCH3* gene variants. Using an AI-based modeling framework and molecular dynamics (MD) simulations^23-25^, we focused on two key NOTCH3 inter-actors, protein O-glucosyltransferase 1 (POGLUT1)^26, 27^, and the antigen-binding fragment (Fab) of an anti-NOTCH3 inhibitory antibody (anti-N3(i) Ab)^28^, to investigate how domain-specific mutations alter protein conformation and interaction dynamics. Simulations of wild-type and mutant proteins in both apo and bound states generated mechanistic hypotheses linking variant-induced molecular perturbations to vascular pathology in patients with *NOTCH3* gene variants. Specifically, disruptions in EGF/POGLUT1 and NRR/Fab interactions may destabilize domain structures and impair their functions, thereby perturbing pathogenic Notch signaling pathways. The observed associations between mutations and neuroimaging markers may be partly explained by mutant proteins exhibiting decreased binding affinities, weakened inter-molecular interactions, and reduced structural stability or functional dynamics. Collectively, our findings elucidate potential mechanisms underlying characteristic neuroimaging features and identify candidate molecular interfaces that may contribute to disease pathogenesis.

## Methods

### Ethics Statement

This study adhered to the principles of the Declaration of Helsinki (1975) and was approved by the local ethics committee of the Third Affiliated Hospital of Sun Yat-sen University with ethical approval numbers: [2011] 2-48 and [2020] 02-148-01. Written informed consent was obtained from each patient involved in the study.

### Study design and participants

Forty genetically verified patients with *NOTCH3* gene variants via the next-generation sequencing procedure were recruited from the Third Affiliated Hospital of Sun Yat-sen University between 2018 and 2023. The symptomatic probands in the pedigrees were included, while the unaffected carriers were excluded. The clinical manifestations and neuroimaging markers were, therefore, used to reveal the correlations with the genetic variants and protein mutations. Clinical information, including demographics (age, sex assigned at birth; sex and ethnicity (Han Chinese) from self-report), neuroimaging markers (white matter hyperintensities (WMHs), microbleeds or macro-bleeds, lacunes, perivascular spaces (PVS), acute cerebral microinfarct (ACMI), and large artery stenosis), vascular risk factors (hypertension, diabetes mellitus, hyperlipidemia, homocysteine (Hcy), smoking, patent foramen ovale (PFO), and atrial fibrillation (AF)), baseline evaluations (Montreal Cognitive Assessment (MoCA), Mini-Mental State Examination (MMSE), and EuroQol 5-Dimension 3-Level questionnaire (EuroQol EQ-5D-3L) scores), and clinical manifestations (migraine, neuropsychological disorder, stroke, and cognitive impairment), were collected and evaluated for each patient as the previous study^29^.

### Binding molecule prediction by an AI-driven structure-based pipeline

The NOTCH3 whole structure was downloaded from AlphaFold Protein Structure Database (entry: Q9UM47). The EGFs, NRR, and Dis locations were obtained from the UniProt Database and extracted from the whole protein. All substructures were searched against the Protein Database (PDB) using Foldseek. The top five complexes with significant sequence-identity, probability, and E-value for each substructure were selected (supplementary Tab. S5). The binding molecules POGLUT1 (entry: 5L0R^30^) and anti-N3(i) Ab (entry: 6XSW^28^) were for wide-type complex structure. All wild-type EGFs/POGLUT1 and NRR/Fab were predicted using AlphaFold3, all structures with high ipTM and pTM scores (supplementary Tab. S4) were for mutant complex structure by Scrwl4.

### MD simulation

For wild-type and mutant proteins, Amber20 and Amber21tools were used for molecular dynamics (MD) simulations in explicit solvent systems, and the force field ff03CMAP, proper for ordered and disordered proteins, was used. All systems were solvated in a truncated octahedron box of TIP3P waters with a buffer of 10 Å, and neutralized by adding counterions. The LEAP module was for assigning hydrogen atoms, and the SHAKE algorithm for bond constraints, including hydrogen atoms. The long-range electrostatic interactions were handled by the Particle Mesh Ewald (PME) method. For detailed MD steps, first, the systems went through 5,000-step steepest descent and 15,000-step conjugate gradient minimizations to remove structural clash. Second, 1 ns heating up and 1 ns equilibration in NVT ensembles were employed. Third, five trajectories of 100 ns each in NPT ensembles were produced, improving configurational sampling and statistical reproducibility. The pressure and temperature were 1 bar and 310 K, respectively. Berendsen barostat and thermostat (coupling constant of 2 ps) were used^31^. The time step was set to 2 fs. The Particle Mesh Ewald Molecular Dynamics (PMEMD) engine of Amber20 was used to perform the steps. Similar protocols were found for analogous systems^32-34^. The stability and convergence were supported by root mean square deviation (RMSD) analysis (supplementary Figs. S2 and S7).

### Conformational and functional Analysis

The last 50 ns of five trajectories, where RMSDs reached the plateau, were concatenated to analyze conformational and functional changes^35, 36^. For structural changes, RMSD quantified global structural deviation between conformations, root mean square fluctuation (RMSF) measured per-atom positional fluctuations and flexibilities, and radius of gyration (RG) characterized overall molecular compactness. For intermolecular interactions and binding affinities, electrostatic and hydrophobic interactions, and hydrogen bonds were calculated, where two nonadjacent residues were in contact if amino acids average distance was less than 6·5 Å, and in electrostatic contact if oppositely charged and less than 11 Å. Solvent accessible surface area (SASA) quantified surface area of molecule exposed to solvent, enabling hydrophobicity assessments upon protein folding. Molecular mechanics generalized born surface area (MMGBSA) of each trajectory estimated binding free energies by combining molecular mechanics energies, implicit solvation, and surface area terms. For collective motions, dynamic cross-correlation matrix (DCCM) identified pairwise correlated c_ij_ regarding similar-correlated (0 to +1), anti-correlated (-1 to 0), or uncorrelated (0) motions between residues i and j, while principal component analysis (PCA) reduced trajectory dimensionality to reveal dominant collective motions driving conformational changes. Protein-protein interactions (PPIs) in the STRING Database characterize how proteins bind and functionally influence each other, enabling critical applications in signaling pathways and disease mechanisms. VMD and Gnuplot were used to plot.

### Statistical analysis

The differences of RMSF, DCCM, and MMGBSA values between wild-type and mutant proteins were calculated. Mann-Whitney U test, a non-parametric statistical method, was used to compare whether two independent samples (e.g., SASA values) originate from the same data distribution.

### Role of the funding source

The funder of the study had no role in data collection, analysis, or interpretation; trial design; patient recruitment; or any aspect pertinent to the study.

## Results

### Characterizing clinical and neuroimaging profiles

It is shown in Tab. S1 that most patients carried the missense variants, except one patient had the insertion variant. Among forty patients, seventeen carried c.C1630T (ID 11), three carried c.G317C, c.C421T, and c.C1759T (IDs 4, 7, and 12), two carried c.T1591C (ID 10), and the remaining carried one. The variants c.C3760T and c.C6442G (IDs 16 and 18) were in one patient. Furthermore, eight variants have not been reported as disease-causing in the literatures of public HGMD (denoted by N=No), while the reported ones were with allele frequency < 0.001 in GnomAD. Most variants were pathogenic in GnomAD or ClinVar, while six variants were not accessible (denoted by NA). The exon positions of gene variants (hg19) are shown in Fig. S1, where exons 3, 4 and 11 occupied more variants.

Tab. S2, Fig. S1 and supplementary Fig. S3 summarize the representative neuroimaging markers for individual patients, and supplementary Tab. S5 lists the demographics, neuroimaging markers, vascular risk factors, baseline evaluations, and symptoms for all patients. The patients with single (IDs 1-2, 9, 15, and 18) and complex protein mutations (IDs. 3-8, 10-14, 16, and 17; ID 8 lacks the image data) were analyzed. WMHs were identified for most patients, except for the one with insertion variant. Microbleeds were found in patients with mutant EGFs 1-3, 8, 13-15, and 26 (IDs 2, 3, 5, 7, 9-12, and 15). Of these, microbleeds with mixed lobar and deep distribution were observed with EGF2 p.V98M and p.C108Y, only deep distribution with EGF1 p.R75Q, EGF3 p.R141C, EGF8 p.R332C, EGF13 p.C531R, and EGF13to14 p.R544C, only lobar with EGF15 p.R587C and EGF26 p.C1022S (Fig. S1 and Fig. S3). The higher numbers of 65, 56, 48, 29, 10, and 9 were with EGF2 p.C108Y, EGF13 p.C531R, EGF8 p.R332C, EGF15 p.R587C, EGF2 p.V98M, and EGF13to14 p.R544C, respectively (Tab. S4).

Except for microbleeds, macro-bleeds in the left external capsule were with EGF15 p.R607C (ID 13), multi-macro-bleeds with mixed lobar and deep distribution were with both EGF32 p.P1254S and Dis p.L2148V (IDs 16 and 18 for one patient). Furthermore, lacunes were found in patients with IDs 2, 3, 5, 9-12, and 15, PVS with IDs 2, 3, and 9-12, and ACMI with IDs 3, 10-12, and 15 (Tab. S4).

In summary, EGF2 p.V98M, p.C108Y, and EGF13 p.C531R (IDs 3, 5, and 10) led to severe microbleeds, while EGF32 p.P1254S and Dis p.L2148V (IDs 16 and 18) led to macro-bleeds. Furthermore, the microbleeds, lacunes, PVS, and ACMI were found for EGF1 p.R75Q, EGF2 p.V98M, EGF13 p.C531R, EGF13to14 p.R544C, and EGF15 p.R587C (IDs 2, 3, and 10-12).

### Predicting binding molecules of NOTCH3 associated with the signaling pathway

Four binding molecules (Jag1, POGLUT1, Soluble Tissue Factor, and anti-NRR Fab) of NOTCH3 were identified by the independent computational pipeline (Tab. 1). The proteins NOTCH1 EGF12 and NOTCH3 EGFs are homologous with a shared POGLUT1-binding motif (C^1^X_a_SX_b_PC^2^), although the direct evidences of NOTCH3/POGLT1 complex were lacking (at that time). As in Figs. 3 and 4, STRING protein-protein interaction database confirmed that NOTCH3 and POGLUT1 were embedded within a highly interconnected network associated with the Notch signaling pathway, including NOTCH3-LFNG (functional score 0.969), POGLUT1-LFNG (0.408), and POGLUT1-F7 (0.916),

**Table 1:**
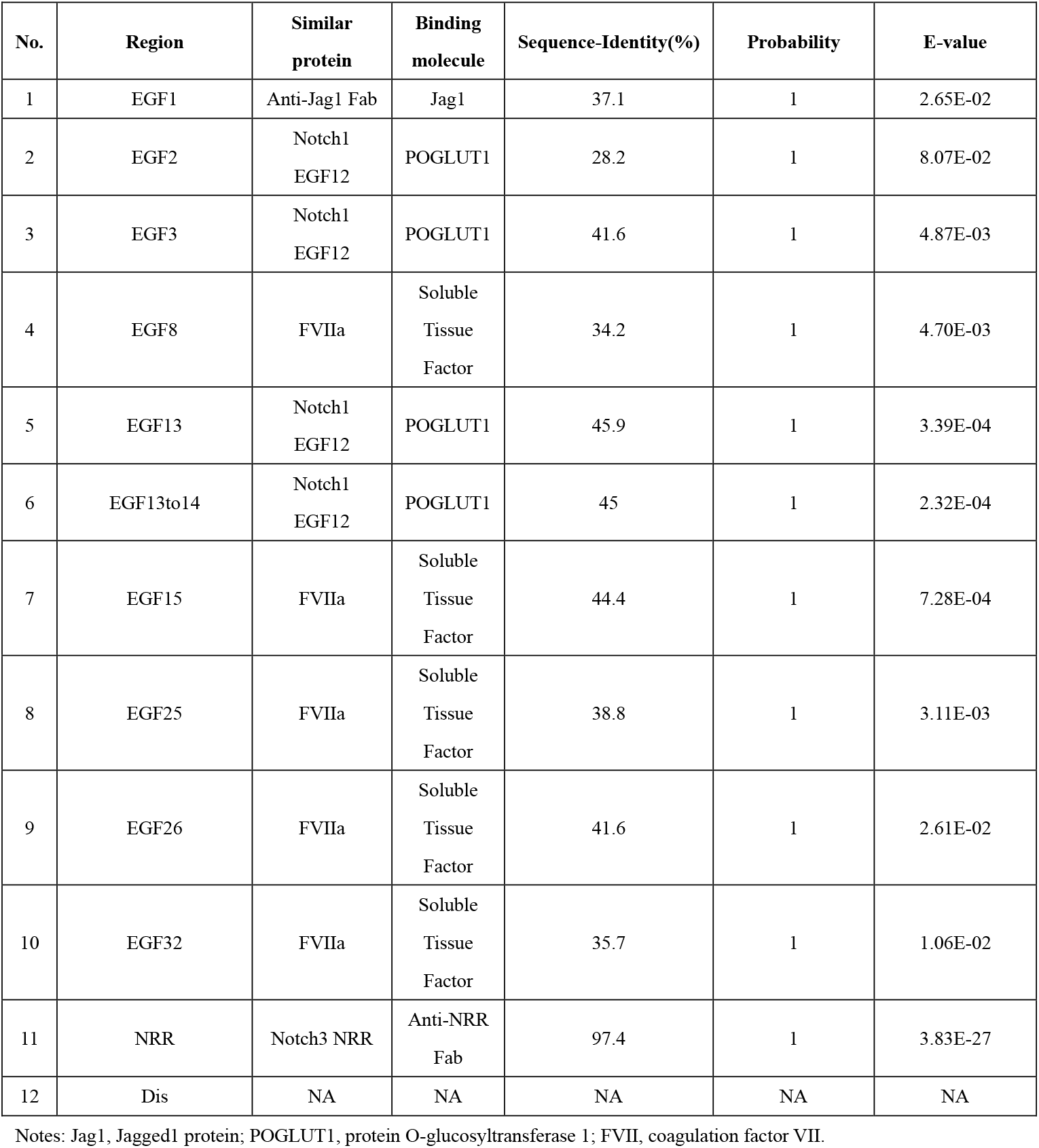
Significant binding molecules for different protein regions of NOTCH3.

where F7 (coagulation factor VII) and NOTCH3 share the conserved POGLUT1-binding motif^30, 37^. Collectively, these results established POGLUT1 as a key candidate interacting molecule of NOTCH3. Based on this finding, we constructed an initial NOTCH3 EGF/POGLUT1 complex model, focusing on EGF repeats containing the conserved POGLUT1-binding motif to elucidate the general mechanism underlying POGLUT1 recognition of NOTCH-family EGF domains. The complex of NOTCH3 NRR/Fab was also constructed.

### Worsening binding affinity and intermolecular interaction

Tab. 2 lists the domain, region, disulfide (SS) bond, and motif of proteins used for structural and functional analysis. Specifically, most mutations were in extracellular domain but the Dis mutation in intracellular. EGFs 2, 3, and 15 contained 4, 2, and 2 variants, and the remaining one variant, respectively. The disrupted SS bonds or binding motifs were in EGF2 pC106S, p.C108Y, and EGF13 p.C531R, or EGF15 p.R587C and EGF32 p.P1254S.

**Table 2:**
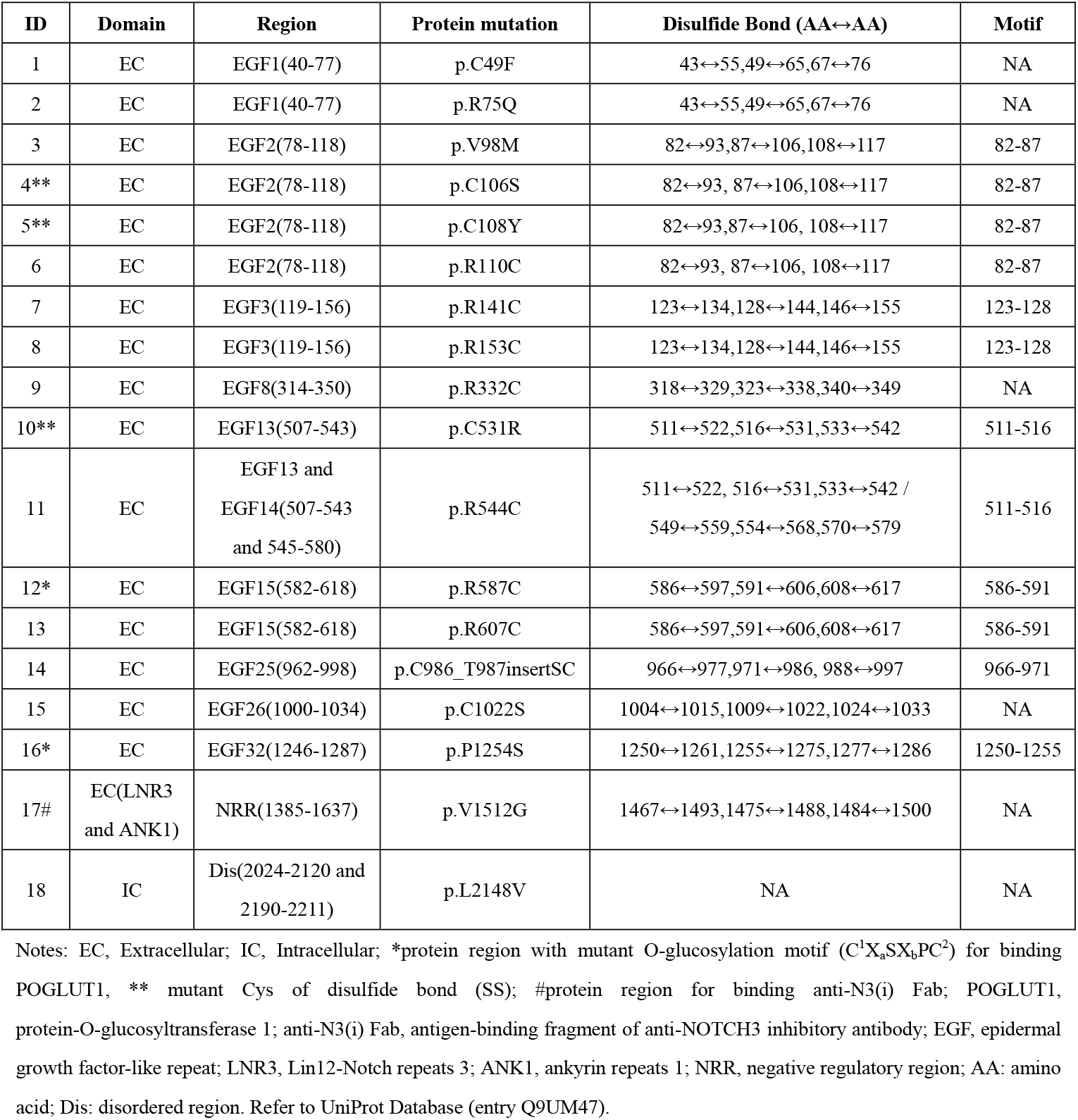
Protein mutations in NOTCH3 domains and regions.

As in Tab. 3 and Fig. 1, for most mutant complexes, the increased (unfavorable) MMGBSA binding energies (differences ΔΔG > 1 kcal/mol) coupled with reduced electrostatic, hydrophobic, and hydrogen bond contributions as well as increased hydrophobic exposure areas were obtained. The larger differences were found for EGF13 C531R, EGF15 R607C, and R587C (18·10, 6·53, and 6·06 kcal/mol, respectively). The number of residue-to-residue inter-molecular interactions decreased significantly for EGF13 C531R (mutant/wild type = 10/90), EGF15 R607C (34/68), and NRR V1512G (164/305), see Fig. 1A and supplementary Tab. S6. The increased solvent accessible surface areas (SASAs) tending to form aggregated structures were for EGF2 C106S, EGF15 R587C and R607C (significant p and Cliff’s δ values), see Fig. 1B. For EGF25 C986iSCiT987, the insertion mutation SC between C986 and T987 reduced the inter-molecular interactions and were weakly related to the hydrophobic exposure areas, while for EGF13to14 R544C, the number increased and the exposure area decreased.

**Table 3:**
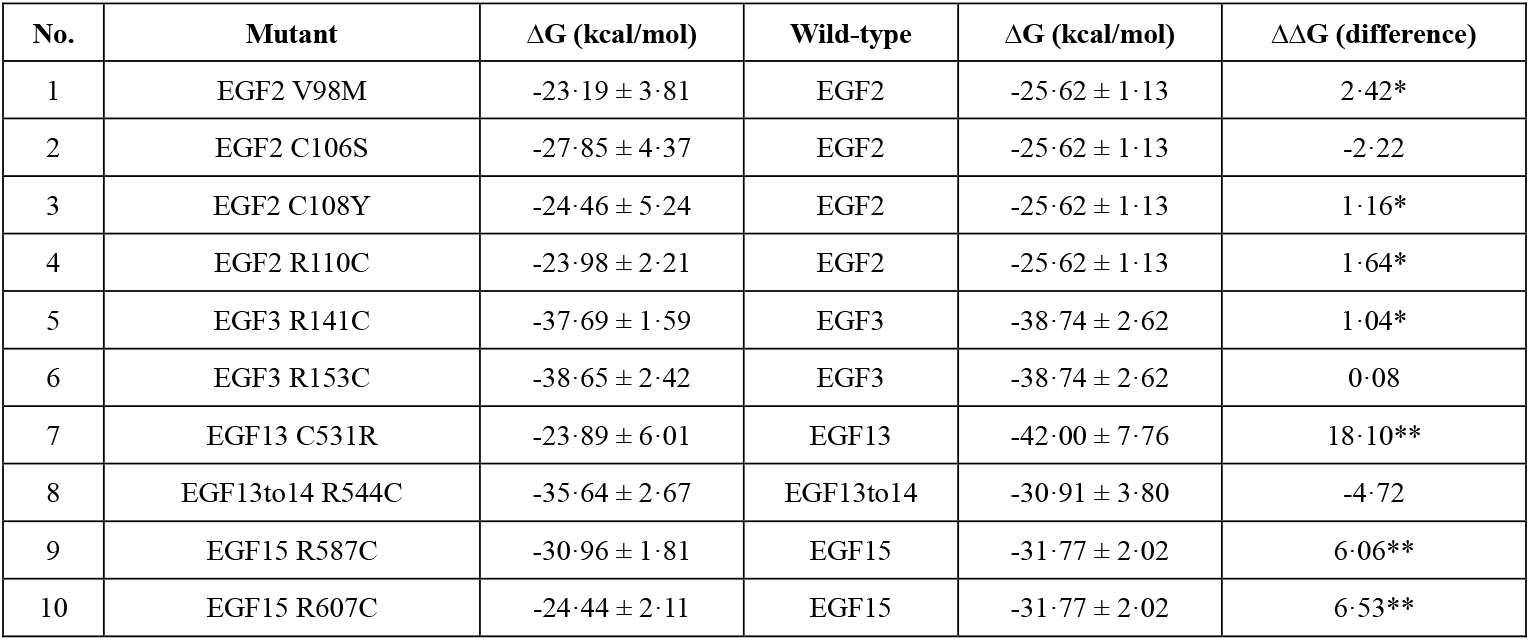
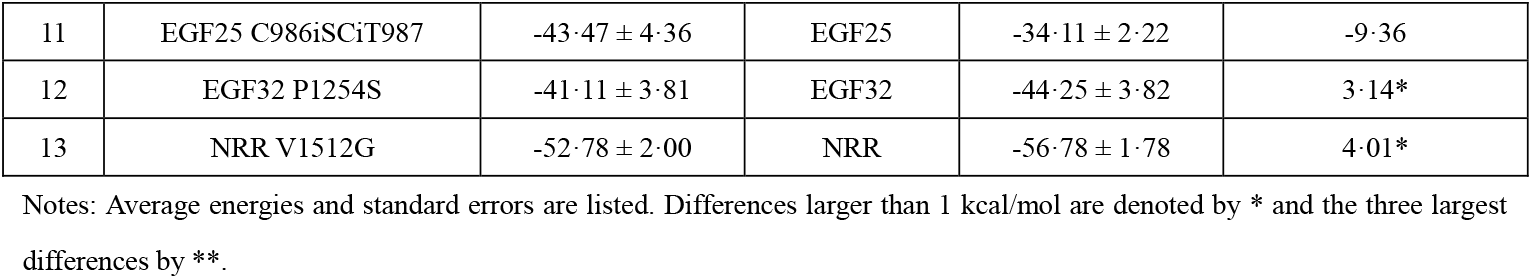
Binding free energies for wild-type and mutant complexes by MMGBSA model.

**Fig. 1:**
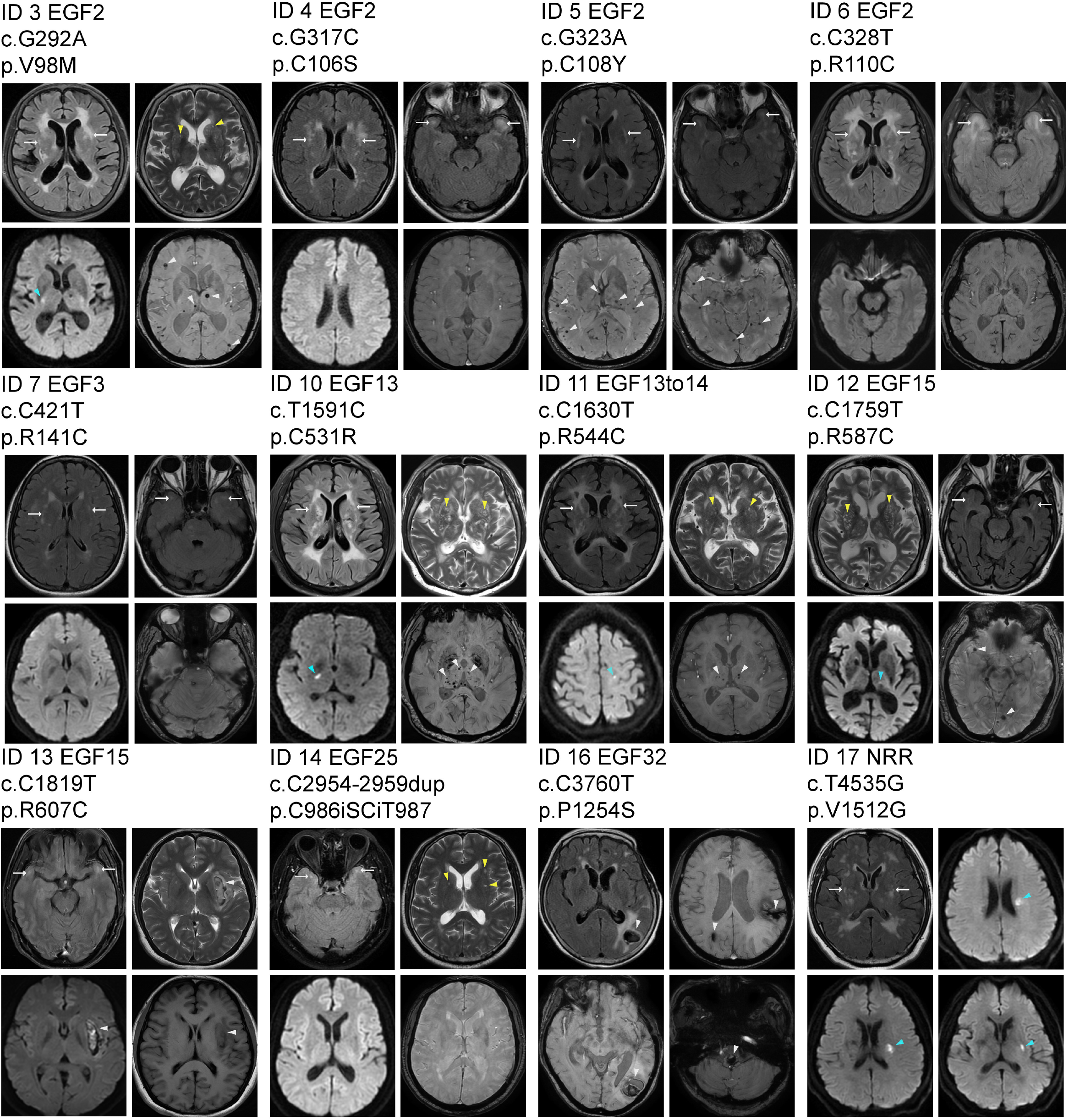
Neuroimaging characteristics for patients carrying NOTCH3 EGFs/POGLUT1 and NRR/Fab gene variants and protein mutations. The characteristics white matter hyperintensities (WHMs, white arrows), microbleeds (white arrowheads), macro-bleeds (white arrowheads), lacune, perivascular spaces (PVS, yellow arrowheads), and acute cerebral microinfarct (ACMI, blue arrowheads) are shown, which were observed on T2FLAIR, SWI, T1FLAIR/T2WI/SWI, T2FLAIR, T2WI, and DWI, respectively. SWI, susceptibility-weighted imaging; DWI, diffusion-weighted imaging.

In summary, for missense mutations in complexes EGFs/POGLUT1, the binding affinities reduced significantly for EGFs 13 and 15, while decreased inter-molecular interactions and increased hydrophobic exposure areas were obvious for EGFs 2, 13, and 15, suggesting weak or ineffective binding of mutants.

### Decreasing conformational stability and compactness

Fig. 4 shows persistently large RMSF, RMSD, and RG values for mutants, which typically indicated significant structural instability or flexibility. As in Figs. 4A and supplementary Fig. S5, the larger RMSF differences between wild-type and mutant EGFs (AA357-398) were revealed for EGFs 2, 3, 13, and 15 (especially for EGF2), while margined differences for others. Furthermore, Fig. 4B shows more expanded RMSD-RG free energy landscapes of low-energy conformations (blue regions) for mutants, where EGF2 C106S, C108Y, and R110C displayed very strong effects, leading to distorted anti-parallel β conformations (Fig. 4C). Interestingly, the narrow landscape of EGF13to14 R544C indicated the aberrant “stable and rigid” structure of two EGFs, where the glycosylation sites were covered (Fig. 4C). This was consistent with the findings for inter-molecular interaction and exposure area in Fig. 3 Refer to supplementary Fig. S6 for all representative structures corresponding to the lowest energy (mutant residues, SS bonds, and glycosylation motifs are displayed).

For apo-EGFs and apo-Dis in single form (supplementary Figs. S10-S11), large RMSF differences for EGF1 C49F and Dis L2148V, and expanded RMSD-RG landscapes with unfavorable structure (reduced SS bonds) for EGF1 C49F are shown. Interestingly, Dis L2148V resulted in small RMSDs and RGs upon binding.

In summary, the local structural changes were obvious in EGFs 2, 3, 13, and 15, among which EGF2 exhibited the most substantial alterations. The conformations were also changed in apo-EGF1 and apo-Dis. Therefore, the mutations resulted in the loss of global compactness and the gain of local flexibility, and hence a net increase in entropy.

### Disrupting collective motions and dominant dynamics

The mutant EGFs/POGLUT1 and NRR/Fab displayed different local and global motions compared to wild-type proteins. As in Fig. 5A and supplementary Fig. S7, the correlation differences indicated that collective residue-to-residue motions were changed, and the effects were more significant for EGFs 2, 3, 13, and 13 to14 (blue and red points). Furthermore, the dominant collective motions (“modes”) of the first and the second components (PC1 and PC2) from PCA analysis driving conformational change are shown. The projection plots of PC1 and PC2 mapped more stable conformational states and fluent transitions for wild-type complexes (supplementary Fig. S8), and porcupine plots visually represented more large-scale motions via long arrows for mutants (Fig. 5B). The negative impacts were more obvious for EGFs 2, 3, and 13 to 14.

For apo-EGFs and apo-Dis (supplementary Figs. S12-S13), the impacts of changing motions were significant for EGF1 C49F and Dis L2148V, where Dis L2148V led to small projections of PC1 and PC2.

In summary, the coordinated residue-to-residue dynamics were disrupted in most mutants, and the dominant protein motions were more disordered and dramatic, especially for EGFs 2, 3, and 13, which agreed well with the previous structural analysis and energy calculations. The influences of apo-Dis were different from others.

### Comparing with published experiments

Previous studies had established NOTCH1 and NOTCH2 as substrates of POGLUT1-mediated O-glucosylation, but whether NOTCH3 underwent the same modification had not been determined at the outset of the study. To address this, we performed computational simulations to characterize the potential interaction between NOTCH3 and POGLUT1. During our analysis, Zhang et al. reported in vitro recombinant expression and glycoproteomic evidence demonstrating that human NOTCH3 is O-glucosylated by POGLUT1^26^. Consistently, Cho et al. subsequently confirmed that mouse NOTCH3 EGFs function as a physiological substrate of POGLUT1, and the complex is required for JAG1- and DLL1-dependent signaling (compared to Figs. 2 and Figs. S2)^38^. These consecutive experimental findings are highly consistent with the NOTCH3/POGLUT1 interaction interface predicted by our computations. The independent validation not only strongly corroborates the reliability of our model for the wild-type complex but also establishes a solid foundation for our core predictions - that pathogenic mutations specifically disrupt this precise molecular interface.

**Fig. 2:**
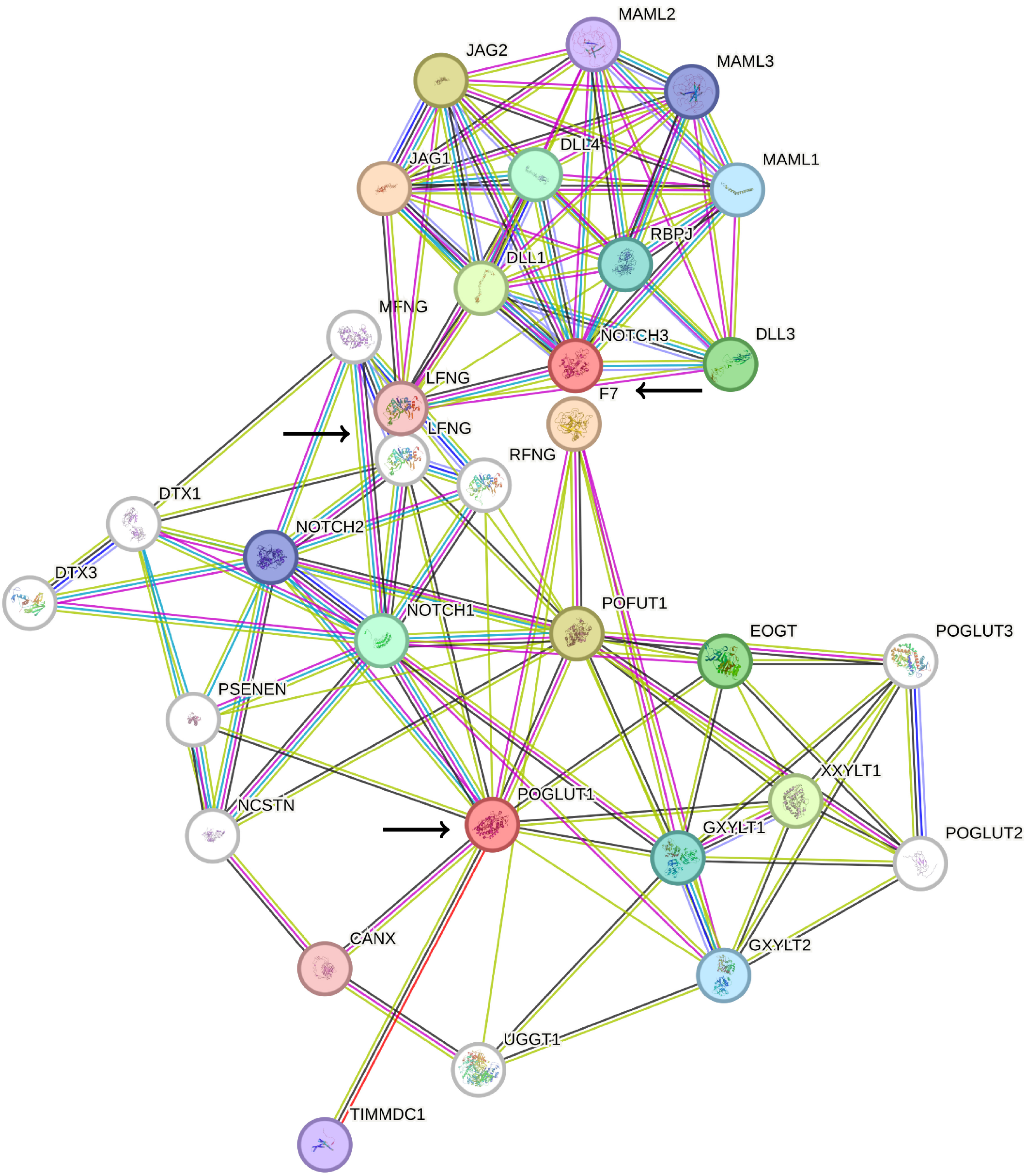
Protein-Protein interactions (PPIs) for signaling pathways of NOTCH3 and POGLUT1 as hub proteins (red nodes).

**Fig. 3:**
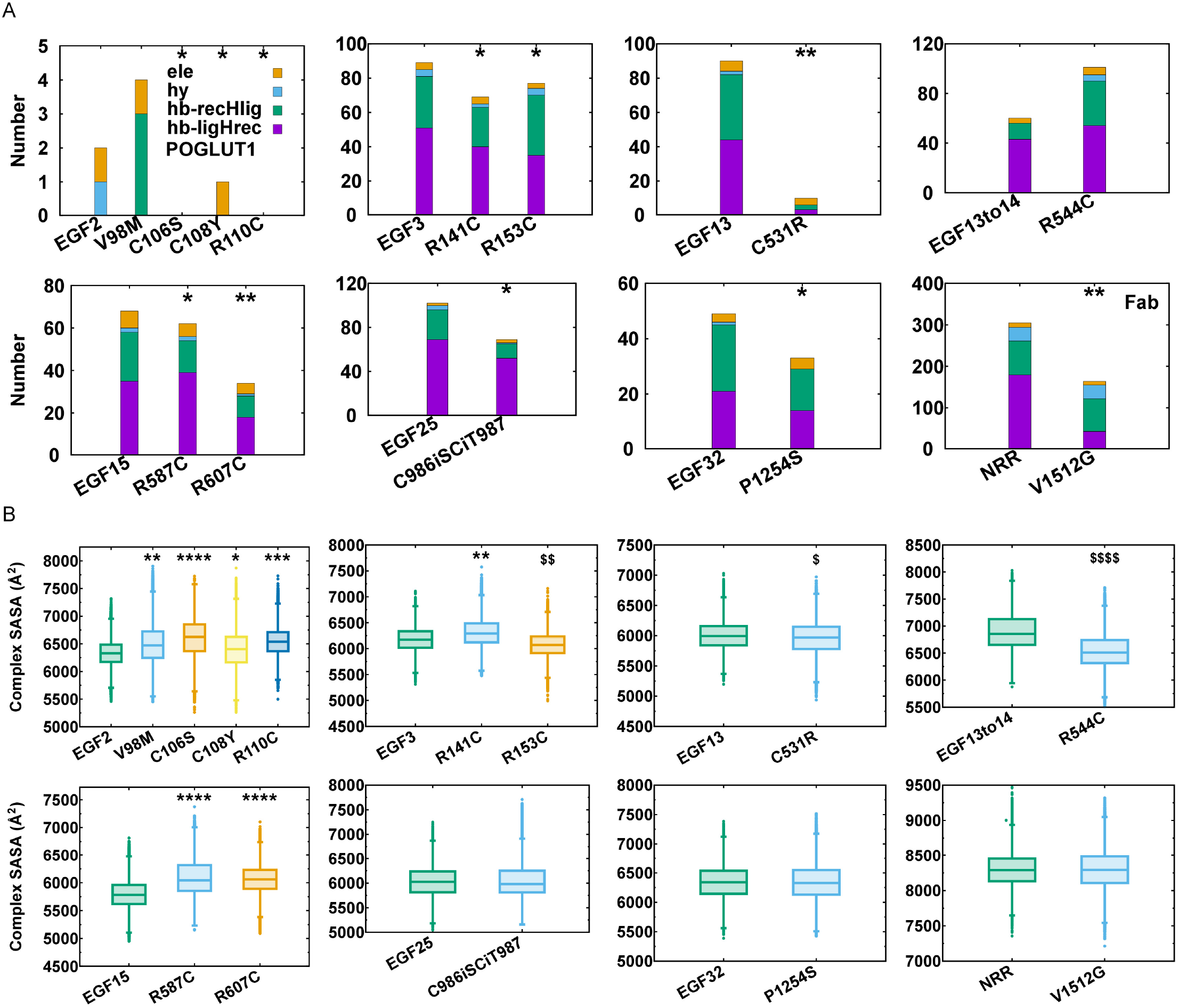
Intermolecular interaction and solvent area exposure. (A) Stable intermolecular interactions including electrostatic contacts (ele), hydrophobic contacts (hy) and hydrogen bonds (hb) where receptors POGLUT1 and Fab as hydrogen donors (hb-recHlig), or ligands EGF and NRR as hydrogen donors (hb-ligHrec); mutants with interactions decrease are denoted by * and the three with the greatest decrease by **. (B) Solvent accessible surface areas (SASAs) of hydrophobic residues of complexes, where p-value ≤ 0·0001 (both * and $), Cliff’s Delta values are -0·147 < δ ≤ 0 (*), -0·33 < δ ≤ -0·147 (**), -0·474 < δ ≤ -0·33 (***), δ ≤ -0·474 (****), 0 < δ ≤ 0·147 ($), 0·147 < δ ≤ 0·33 ($$), 0·33 < δ ≤ 0·474 ($$$), δ ≥ 0·474 ($$$$), mutant with SASA larger (*) or smaller ($) than wild-type.

**Fig. 4:**
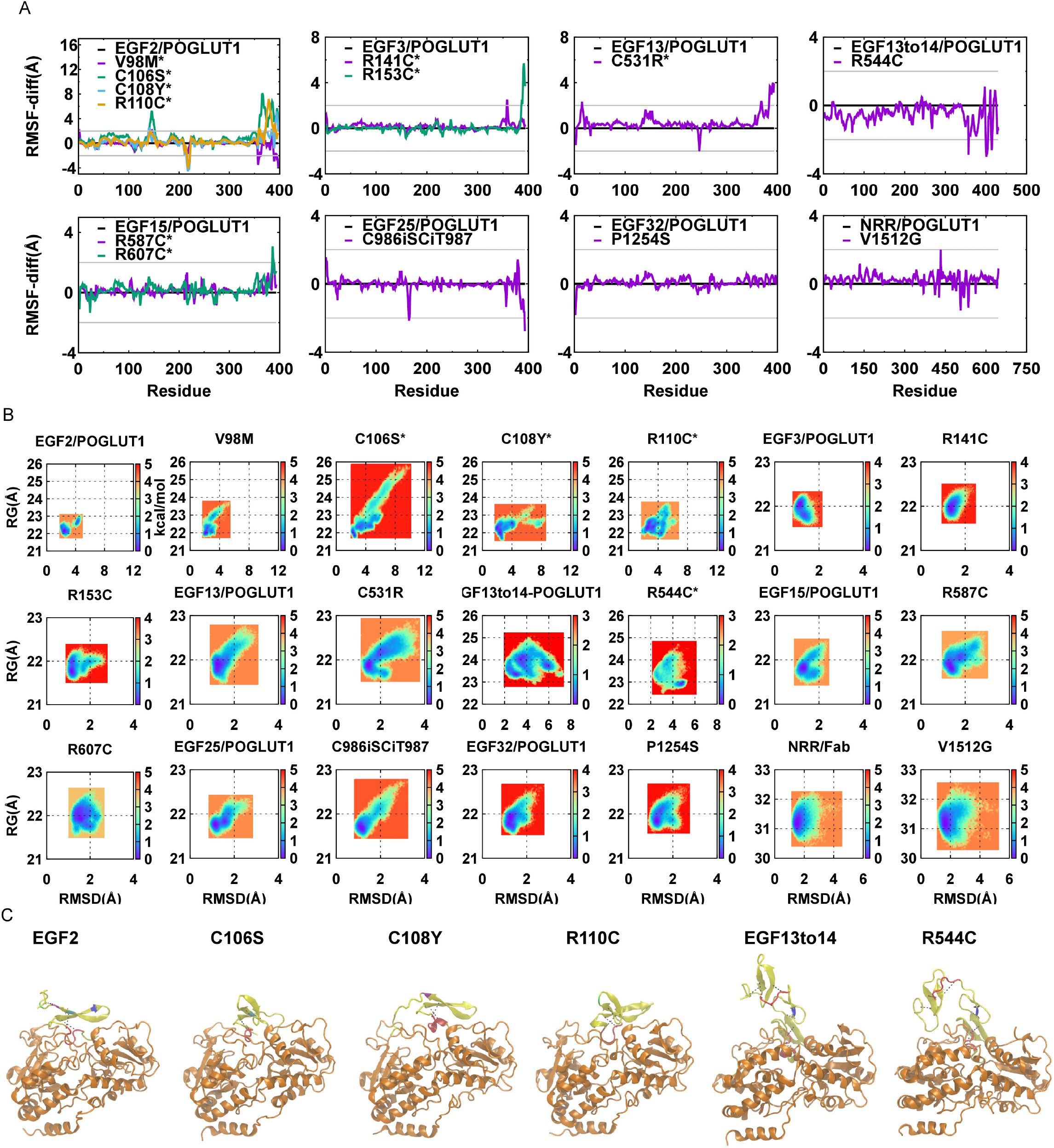
Conformational stability and compactness. (A) Differences of root mean square fluctuation (RMSF) values for wild-type and mutant complexes; (B) Free energy landscapes of root mean square deviation (RMSD) and gyration radius (RG) for wild-type and mutant complexes. (C) Stable conformations corresponding to minimal energies in RMSD-RG landscapes, where EGFs are colored in yellow (top), POGLUT1 in orange (bottom), the glycosylation motifs in red, the mutant and wild-type residues in other colors, the disulfide bonds are represented by dashed dots. Differences larger than 2 Å are denoted by *.

**Fig. 5:**
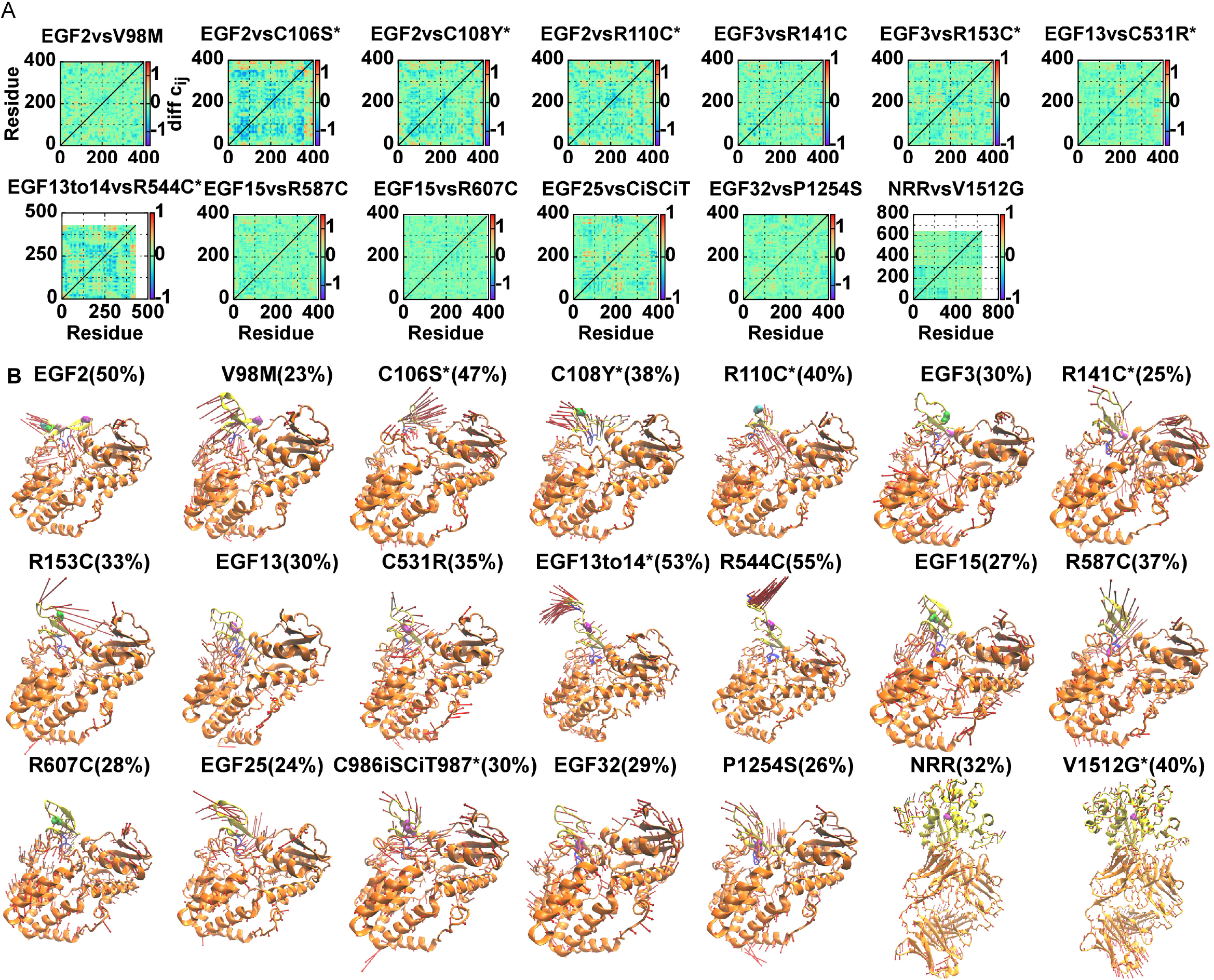
Collective motions and dominant dynamics. (A) Differences of dynamic cross-correlation matrix (DCCM) values for wild-type and mutant complexes. (B) Dominant motions along the first components (PC1) with contribution percentages of the principal component analysis (PCA) for wild-type and mutant complexes, where EGFs and NRR are colored in yellow (top), POGLUT1 and Fab in orange (bottom), the mutant and wild-type residues in other colors. Large differences are denoted by *.

Furthermore, the previous assay shows that cysteine residues and disulfide bonds in EGFs1-3 are essential for maintaining normal NOTCH3 structure, as mutations affecting cysteines by loss, modification, or gain (e.g., R141C) result in structural abnormalities and only R75C/P/G induce structural shifts^39^. R141C also leads to the protein multimerization and the high disease severity^40, 41^.

These findings are consistent with our calculations, for example, R75Q displayed a marginal difference in structural and functional changes compared to wild type. The experiment of all non-cysteine substitutions at residue 49 (e.g., C49F) reveals that the mutations result in the significant conformational changes^42^, which is also in good agreement with our computations. Refer to the supplementary Figs. S7-S11 for C49F and R75Q.

## Discussion

Our study highlights the potential of the contemporary strategy of AI calculations and MD simulations for the systematic evaluations of the NOTCH3 variants. For NOTCH3 variants, two important binding molecules, POGLUT1 and anti-N3(i) Ab, related to the pathogenetic signaling pathways have been identified. Importantly, the binding partner POGLUT1 was determined by both the computational and the very recent experimental studies. The effects of the NOTCH3 variants depend on the locations (e.g., in EGFs 1-6), the types (e.g., forming the disulfide bonds), and the structures and functions (e.g., containing the glycosylation motif). The effects of some specific mutations in single forms are consistent with previous studies. Targeting POGLUT1 to modulate EGF-like domains and using the Fab region to stabilize NRR complexes represents a promising therapeutic approach deserving rigorous exploration, which could be transformed into the future development of disease-modifying therapeutics for NOTCH3 variants.

The research on the post-translational processing, including glycosylation of NOTCH3 variants, attracts more attention while the current investigations are still limited^17, 40, 43^. For example, the mutations impairing Fringe-mediated O-fucose elongation on NOTCH3 EGFs 1-5 have been reported^44^, the mutant protein aggregation propensity may result from the lunatic fringe^45^, and the signaling activity and turnover of mutants decrease with increasing Radical fringe (RFNG) expression^46^. Furthermore, the recent study shows that the NRR plays a central role in signaling activation by directly interacting with multiple Jagged1 domains, forming a nonlinear interaction network that redefines the Notch1-Jagged1 activation mechanism^47^. Our investigations of POGLUT1 and NRR serve as the promising basis for future researches.

This study has several limitations. First, the patient sample size and number of variants are relatively small; larger multi-center cohorts will be necessary to validate our findings. Second, neuroimaging assesses neural activity only indirectly and is constrained by limited spatial and temporal resolution, as well as by participant age and medication status, which together restrict causal interpretation. Third, experimental validation remains limited, while the AI and MD predictions are powerful and informative, they should be considered as hypothesis-generating until confirmed by independent wet-lab studies.

Our study links the gene variants and protein mutations to clinical characteristics and neuronal phenotypes, which enhances the understanding of the molecular pathogenesis of CADASIL. Targeting POGLUT1 to modulate EGF-like domains and using the Fab region to stabilize NRR complexes represents a promising therapeutic approach deserving rigorous exploration.

## Data Availability

Data are available upon reasonable request. The clinical, genetic, and computational data that support the findings of the study are available from the corresponding authors, Dr. Wei Qiu, Dr. Zhengqi Lu, and Dr. Qingfen Yu, upon reasonable request. There is no additional information available.

## Acknowledgments

We express our deepest appreciation to all patients and their family members for the participation and support of our study. We gratefully thank the staffs at Mental and Neurological Diseases Research Center and Center for Big Data and Artificial Intelligence of The Third Affiliated Hospital of Sun Yat-sen University for providing the clinical and computational resources.

## Study Funding

The funding provided by Hundred Talents Program of Sun Yat-sen University (P000-2105) is gratefully acknowledged.

## Contributors

**Xuejiao Men:** conceptualization, data curation, formal analysis, investigation, resources, project administration, resources, supervision, validation, visualization, writing - original draft, writing - review & editing. **Li Zhang:** data curation, formal analysis, investigation, software, visualization (MD simulations). **Sanxin Liu**: data curation, formal analysis, writing - original draft, writing - review & editing. **Shunzhou Wan:** methodology, validation, writing - review & editing. **Wei Qiu:** resources, project administration, supervision, writing - review & editing. **Zhengqi Lu:** resources, project administration, supervision, validation, writing - review & editing. **Qingfen Yu:** conceptualization, data curation, formal analysis, funding acquisition, investigation, methodology, project administration, resources, software, supervision, validation, visualization, writing - original draft, writing - review & editing.

## Declaration of interests

None declared.

## Declaration of generative AI and AI-assisted technologies in the manuscript preparation process

During the preparation of this work the author(s) used DeepSeek-V3.1 and GPT-5 to improve language and readability. After using this tool/service, the author(s) reviewed and edited the content as needed and take(s) full responsibility for the content of the published article.

## Notes

### Competing Interest Statement

The authors have declared no competing interest.

